# A pilot study of exercise training for children and adolescents with inflammatory bowel disease: an evaluation of feasibility, safety, satisfaction, and efficacy

**DOI:** 10.1101/2022.10.24.22279788

**Authors:** Mila Bjelica, Rachel G. Walker, Joyce Obeid, Robert M. Issenman, Brian W. Timmons

## Abstract

**Background:** Children with inflammatory bowel disease (IBD) experience extra-intestinal side effects including altered body composition, impaired muscle strength and aerobic capacity. Exercise training may remedy these issues.

**Purpose:** To assess the feasibility, safety, participant satisfaction and efficacy of a training program for youth with IBD.

**Methods:** Children with IBD completed 16 weeks of training (2 supervised+1 home sessions per week). Feasibility was assessed by tracking recruitment, adherence, and compliance rates. Safety was assessed by tracking symptoms and adverse events. Post-training interviews gauged satisfaction. Circulating inflammatory markers, body composition, muscle strength, aerobic fitness, and habitual physical activity, were measured at baseline, mid-training (8 weeks), and post-training.

**Results:** Eleven youth were recruited and 10 completed the study. Participants adhered to 28±1 of 32 prescribed supervised sessions and 8±4 of 16 prescribed home sessions. There were no adverse events, and overall feedback on training was positive. Post-training, we observed an increase in lean mass (+2.4±1.1 kg), bone density (+0.0124±0.015 g•cm^-2^), aerobic fitness (+2.8±5.7 mL•kgLM^-1^•min^-1^), and vigorous physical activity levels (+13.09± 8.95 min•hr^-1^) but no change in inflammation or muscle strength.

**Conclusion:** Supervised exercise training is feasible, safe, and effective for youth with IBD and should be encouraged.

## Introduction

The direct economic burden of inflammatory bowel disease (IBD) in 2018 was estimated to be $2.6 billion in Canada alone.^1^ Rates of IBD continue to rise globally; however, over the last 20 years there has been shift in affected populations. In fact, there is an increasing incidence in pediatric IBD cases compared to cases in adults and the elderly.^2^ In Canada, rates of pediatric IBD are among the highest worldwide, with approximately 600-650 new diagnoses per year among those less than 16 years of age(1).

Children with IBD suffer from numerous extra-intestinal side effects including increased systemic inflammation, altered body composition (i.e., reduced muscle and bone mass, increased fat mass), reduced muscle strength and impaired aerobic capacity, even when in remission.^3–5^ These side effects can be attributed to complex and interconnected factors including disease pathology, medication usage, and physical inactivity.^6^ Together, the inherent and incidental factors associated with pediatric IBD may contribute to the development of a vicious cycle of deterioration in overall health and quality of life. ^3–6^ As such, it is critical to establish strategies to break this feedforward cycle.

Exercise is a simple and accessible intervention with the potential to attenuate some of the extra-intestinal health deficits associated with IBD. In adults with IBD, exercise training can increase muscle strength, lean mass, and bone mass, reduce body fat and improve aerobic fitness, without indications of disease and/or symptom exacerbation.^7–10^ Presently, pediatric IBD exercise training studies have been limited to exergaming interventions and report improvements in exercise capacity, self-reported physical activity, and circulating inflammatory markers.^11,12^ However, no studies to date have assessed the effects of exercise training on body composition, muscle strength, aerobic fitness, or device-based measures of physical activity in children with IBD, nor reported the safety and feasibility of an exercise intervention. Given that children with IBD are less physically active than healthy counterparts,^13^ a disease-appropriate exercise training program using both aerobic and resistance training could be an effective way to engage these youth in physical activity, while also to managing the side effects of IBD.

In order to effectively design a larger, randomized controlled trial, we first opted to perform a pilot study. Therefore, the primary aims of this study were to evaluate the: 1) feasibility 2) safety, and 3) participant satisfaction in an exercise training program for children and adolescents with IBD. The secondary objectives of this study were to assess the physiological effects of a 16-week exercise training program on: 1) body composition, 2) systemic inflammation, 3) muscle strength, 4) aerobic fitness, and 5) habitual physical activity levels in children and adolescents with IBD.

## Methods

### Participant recruitment

Children and adolescents with IBD between 10 to 17 years of age were recruited from the Centre for Child and Youth Digestive Health at McMaster Children’s Hospital, Hamilton, Canada. Eligible patients were in remission (score of >10) according to the pediatric Crohn’s disease activity index (PCDAI) or the ulcerative colitis activity index (PUCAI) and/or physician confirmation. Patients were excluded based on physician recommendation or if they regularly engaged in resistance training ≥ 3 times a week. Informed assent and consent were collected from participants and parents or guardians, as appropriate. This study was approved by the Hamilton Integrated Research Ethics Board.

### Study overview

The 16-week training program consisted of 3 exercise sessions per week: two supervised sessions at McMaster University and one home-based session. Heart rate was monitored at supervised and home sessions. Each session lasted 30-60 min and included both aerobic and resistance exercises. (full details available in **Supplementary Material 2, Supplementary Table 1**). After each training session, children were given a high protein beverage (0.2 g protein/kg body weight) to promote muscle anabolism. Due to dietary allergies, two participants were given an alternative beverage matched for protein content.^14^ Over the course of the study, participants attended 3 assessment visits: 8 days prior to starting the training (PRE), after 8 weeks of training (MID), and 8 days after completing the last training session (POST) to evaluate study outcomes (**Supplementary Figure 1**). Each assessment visit included measures of height, weight, body composition, a fasting blood sample for assessment of inflammatory markers (circulating IL-6 and TNF-α), muscle strength, aerobic fitness, and 7-day habitual physical activity. One-repetition maximum (1RM) and maximum repetitions in 45 sec was evaluated after PRE and MID assessment visits to standardize training intensity (**Supplementary Material 1**). During their POST assessment, participants engaged in a structured qualitative interview with the investigator to evaluate participant satisfaction with the training study.

### Primary outcomes

#### Feasibility

Feasibility included measures of recruitment, retention, adherence and compliance rates. Recruitment and withdrawal rates were tracked over a 9-month period to assess the feasibility of enrollment and retention at a single site. Based on the rates of absences in an adult IBD exercise training study with a similar training frequency, we defined our training program to be feasible if participants on average could adhere to 80% of the prescribed exercise sessions.^15^ Participant adherence to the exercise training program was assessed by the number of exercise sessions completed relative to total prescribed sessions. For supervised sessions, attendance and reasons for absences were tracked. Home sessions were marked as completed when there was evidence of HR monitor wear for exercise. Compliance was defined as the proportion of each training session that was completed as prescribed. To assess participant compliance to supervised sessions, training logs were maintained for each participant, tracking the prescribed exercise as well as the completed intensities and repetitions or duration of each workout. Compliance was reported as the percent of supervised sessions where participants completed 100% of the prescribed exercise without any modifications to exercise intensity, duration, or repetitions, as well as how many sessions required an alteration in resistance or aerobic training volume.

#### Safety

Safety was assessed by tracking changes in IBD-related symptoms bi-weekly, using a participant-completed questionnaire. Adverse events were also tracked over the course of the study. Adverse events were defined as any event that occurs during the course of exercise training that causes the participant physical or psychological harm, such as musculoskeletal injuries or hospitalizations. An exit criterion was defined as any participant who experienced a disease flare-up that required hospitalization and initiation of steroids.

#### Participant satisfaction

Participants engaged in a qualitative interview with an investigator (RW) post-training to evaluate their level of satisfaction with the exercise training program and identify areas for improvement. The interview was audio-recorded, transcribed by RW, and participant answers were extracted and reported as frequencies by MB. Participants were asked the following specific questions:

1. Which parts of the training program did you enjoy and what did you like about them?
2. What parts of the training program did you not enjoy and what about them did you not like?
3. Which parts of the training program would you recommend improving and how should we go about making these improvements?

### Secondary outcomes

All secondary outcomes were assessed at PRE, MID, and POST. Detailed descriptions of each outcome are provided in the **Supplementary material 1**. In brief, body composition was assessed by dual energy x-ray absorptiometry (DXA; Hologic, Marlborough, Massachusetts). High-sensitivity enzyme-linked immunosorbent assays were used to quantify circulating levels of interleukin-6 and tumor necrosis factor alpha (RND Systems; Minneapolis, Minnesota) from a fasting blood sample (10 hours). Muscle strength (i.e., isometric and isokinetic at 60,120,180°•s^-1^ for elbows and knees) was assessed on a Biodex Isokinetic Dynamometer (System Pro 4; Biodex Medical Systems Inc, Shirley, New York), while aerobic fitness was measured using a cardiopulmonary exercise test on a stationary cycle ergometer (Corival, Lode; The Netherlands). Free-living physical activity was assessed by 7 days of accelerometry (ActiGraph GT1M; ActiGraph Corp, Pensacola, FL). **Statistical analysis:**

Descriptive statistics were used to characterize feasibility, safety, and satisfaction outcomes. All secondary outcomes (physiological data) were assessed for normality using the Shapiro-Wilk test. PRE, MID, and POST outcomes were compared using one-way repeated-measures ANOVAs for normally distributed data and Friedman’s ANOVAs for non-normally distributed data. Post hoc analyses were completed using pairwise comparisons with a Bonferroni correction or Wilcoxon signed-rank test with a Bonferroni correction for normally and non-normally distributed data, respectively. Statistical significance was set at p ≤ 0.05. Effect sizes were calculated as Cohen’s *d* for changes in body composition, muscle strength and aerobic fitness variables from PRE to POST.^16^ A correction factor was used to adjust for our small sample size.^17^ The correction factor was calculated according to the following equation (Eq1.):

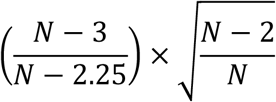

Eq1. Small sample size correction factor

An effect size of 0.2 was defined as small, 0.5 as medium and 0.8 as large.^18^ Given that this was a pilot study, sample size was not calculated a priori.^19^

## Results

### Recruitment and retention

Participant recruitment began in May 2013 and ended February 2014, and participant training began July 2013 and ended June 2014. In this time, a total of 217 patients were screened for eligibility, 93 (42.9% of total) patients were approached, and 11 (11.8% of approached) consented to participate. Ten (90.9% of enrolled) patients completed the 16-week exercise training program, and one dropped out for medical reasons unrelated to the study (**Figure 1**).

**Figure 1.**
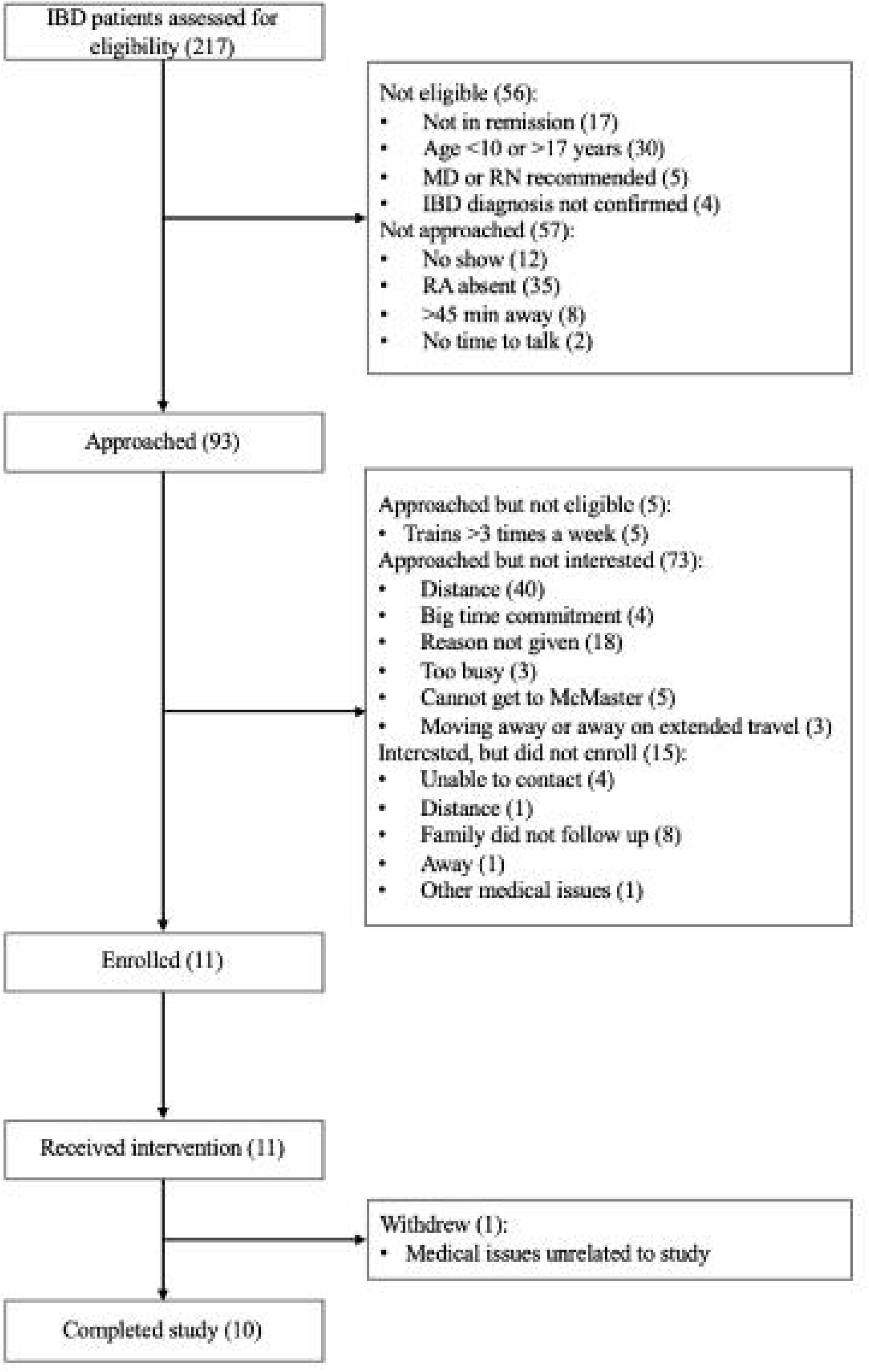
Recruitment process flow chart.

### Adherence

Of the 48 exercise training sessions prescribed, participants completed an average ± SD (range) of 37 ± 5 (range: 32-47) sessions, slightly below our 80% attendance threshold for feasibility. Participants completed 28 ± 1 (range: 26-31) of the 32 prescribed supervised training sessions and 8 ± 4 (range: 4-16) of the 16 prescribed home training sessions. Reasons given for missing supervised training sessions included illness, inclement weather, travel away from home, scheduling conflicts and on one instance, a car accident. Reasons given for missing home training included illness, fatigue, travel away from home, and time constraints.

### Compliance

Participants completed an average of 83 ± 6% (range: 69-92%) of the prescribed exercise at supervised sessions without any modifications. Resistance exercises were modified at 3 visits or 12 ± 6% (range: 3-26%) of attended sessions by reducing the intensity or repetitions of at least one set of exercise. Reasons for modifying resistance exercises included: the resistance was too difficult, and joint pain (i.e., repetitions and/or intensity of leg curls were reduced due to knee pain). Aerobic exercise was modified during 1 visit or 5 ± 4% (range: 0-13%) of sessions by reducing duration. Reasons for modifying aerobic exercise sessions included: participants came in too tired from previous activities (i.e., soccer try-outs, gym class), participants felt the exercise was too difficult to complete, participants reported joint pain, or participants felt unwell that day due to disease symptoms. In two instances participants did not complete the final set of resistance training and/or steady state cycling because they had to leave the session early due to prior commitments. Home compliance was not assessed because the heart rate monitor only provided overall session averages, and participants did not regularly log their exercise sessions.

### Safety

During the intervention, two participants experienced minor disease flares that resolved without the need for medical intervention. No adverse events associated with the exercise training were reported. Based on results from the bi-weekly symptom questionnaire, self-reported general well-being 4 of 10 participants reported themselves as “well” from beginning to end (the highest rating on this scale). Three participants fluctuated between “well” and “slightly below par” throughout the study, but ended the study feeling “well”. Two children finished with “well” despite starting with “poor” and “slightly below par”, while one child started with “well” and finished with “slightly below par”. Complete results for all items in the questionnaire can be found in **Supplementary Tables 3-15. Participant satisfaction:** Nine of 10 participants completed the satisfaction interview. Participants likes (10 unique responses from 9 participants) and dislikes (5 unique responses from 8 participants) are reported by frequency in **Supplementary table 2**. Resistance training activities performed on machines received the most frequent positive feedback. One child said they felt uncomfortable with their body and self-conscious exercising in public places, but they enjoyed the opportunity to exercise one-on-one with a trainer. Another child indicated they felt more motivated to exercise in the presence of a trainer, unlike at home. Conversely, one participant said they enjoyed the structured home sessions because it was more convenient and helped them realize they can be active on their own, at home. The most frequently reported dislike was cycling activities: two participants reported disliking interval cycling, two participants disliked continuous cycling, one participant disliked the aerobic fitness test, and two participants stated that cycling did not feel satisfying because it was done on a stationary bike. In addition, two participants disliked leg extensions because of the pressure they felt on their knees and thighs, one participant disliked home sessions because they were difficult to remember to complete. Finally, participants also provided recommendations on how they would improve the training program. The most notable suggestions were to:

- Create a personalized training goal for each participant (e.g., to be able to perform a chin up by the end of the program).
- More options for aerobic exercises (e.g., ellipticals, treadmills).
- Host group training sessions with multiple children of a similar age.
- Include more parental involvement in the program, so that they can motivate participants to complete home-based sessions.
- Include a follow-up period post-study completion to ensure participants are still training and/or to have a longer training period until the participant is comfortable enough to train on their own.

### Secondary outcomes

#### Participant characteristics

Participant characteristics are displayed in **Table 1**.

**Table 1.** Participant characteristics

| **Mean ± SD (min-max)** | **PRE** | **MID** | **POST** |
| --- | --- | --- | --- |
| N (M/F) | 10 (9/1) | 10 (9/1) | 10 (9/1) |
| CD/UC | 6/4 | 6/4 | 6/4 |
| Age (years) | 15.4 ± 1.2<br>(14.1 – 17.3) | 15.59 ± 1.2<br>(14.2 – 17.5) | 15.8 ± 1.2<br>(14.4 – 17.7) |
| YPHV | -1.3 ± 1.3<br>(-3.2 – +1.6) | -1.2 ± 1.2<br>(-3.0 – +1.4) | -1.1 ± 1.2<br>(-2.9 – +1.5) |
| Tanner (# per stage) |  |  |  |
| II | 1 | 1 | 1 |
| III | 3 | 2 | 1 |
| IV | 4 | 5 | 5 |
| V | 2 | 2 | 3 |
| Height (m) | 1.67 ± 0.72<br>(1.58 – 1.80) | 1.68 ± 0.70<br>(1.58 – 1.81) | 1.69 ± 0.70<br>(1.58 – 1.81) |
| Weight (kg) | 54.3 ± 8.3<br>(42.1 – 68.9) | 55.9 ± 9.1<br>(43.1 – 70.9) | 57.1 ± 9.0<br>(43.1 – 69.9) |
| BMI (kg/m<sup>2</sup>) | 19.4 ± 2.7<br>(15.3 – 24.0) | 19.7 ± 3.0<br>(15.6 – 24.8) | 20 ± 2.9<br>(16.2 – 25.3) |

Data are presented as frequencies, mean ± standard deviation, and range. CD= Crohn's disease, UC= Ulcerative colitis, YPHV= Years to peak height velocity

As expected in children with IBD, the majority of our participants presented with below average height, weight and BMI percentiles.^20^

### Body composition

There were significant increases in lean body mass (LBM) in kg from PRE to MID, MID to POST, and PRE to POST. There was a main effect for training suggesting an increase in LBM% (post-hoc comparisons not significant, NS). Additionally, there was an increase in bone mineral content (BMC) in kg from PRE to MID and PRE to POST. While an effect for training suggests BMD (post-hoc NS) increased over the course of 16 weeks. While we did not observe any change in FBM (kg), FBM% decreased (post-hoc NS). There was also a significant decrease in trunk fat% from PRE to MID (**Table 2**).

**Table 2.** Participant body composition at PRE, MID and POST training

|  | PRE | MID | POST | Effect size | ANOVA or Freidman Statistics |
| --- | --- | --- | --- | --- | --- |
| **BMC (kg)** | 1.74 ± 0.24 | 1.78 ± 0.25* | 1.83 ± 0.26*<sup>§</sup> | 0.29 | Greenhouse Geisser correction used, $\epsilon=0.626$; $F(1.252,11.266)=16.825$ $p<0.05$ |
| **FBM (kg)** | 12.7 ± 5.04 | 12.95 ± 5.46 | 12.8 ± 5.63 | 0.02 | Nonparametric Friedman test, $\chi^2(2)=0.800$, $p>0.05$ |
| **LBM (kg)** | 40.8 ± 5.2 | 42.3 ± 5.3* | 43.2 ± 5.2* | 0.37 | $F(2,18)=2.113$, $p>0.05$ |
| **BMC %** | 3.2 ± 0.4 | 3.2 ± 0.4 | 3.2 ± 0.4 | 0.03 | $F(2,18)=0.988$, $p>0.05$ |
| **FBM %** | 22.5 ± 6.5<sup>+</sup> | 22.1 ± 6.6<sup>+</sup> | 21.5 ± 6.8<sup>+</sup> | -0.12 | Greenhouse Geisser correction used, $\epsilon=0.626$; $F(1.251,11.260)=5.637$ $p<0.05$ |
| **LBM %** | 74.3 ± 6.3<sup>+€</sup> | 74.8 ± 6.3<sup>+€</sup> | 75.3 ± 6.5<sup>+€</sup> | 0.13 | Nonparametric Friedman test, $\chi^2(2)=7.200$ $p<0.05$ |
| **BMD (g·cm<sup>-2</sup>)** | 0.967 ± 0.071<sup>+</sup> | 0.971 ± 0.074<sup>+</sup> | 0.979 ± 0.072<sup>+</sup> | 0.14 | $F(2,18)=3.714$ $p<0.05$ |
| **Trunk fat (kg)** | 4.46 ± 1.86 | 4.42 ± 1.97 | 4.43 ± 2.09 | -0.01 | Nonparametric Friedman test, $\chi^2(2)=0.200$, $p>0.05$ |
| **Trunk fat %** | 18.0 ± 5.2 | 17.2 ± 4.6* | 17.0 ± 5.6 | -0.15 | $F(2,18)=3.958$ $p<0.05$ |

PRE, MID, and POST outcomes reported as mean ± standard deviation. BMC= bone mineral content, FBM= fat body mass, LBM= lean body mass. \* $P<0.05$ compared to PRE, <sup>§</sup> $P<0.05$ compared to MID, <sup>+</sup>Post-hoc test not sensitive enough to detect differences, <sup>€</sup>For non-parametric Freidman test the Bonferroni correction used for post-hoc testing resulted in significance level of $p\leq 0.017$

### Cytokine levels

There were no observed trends for circulating levels of IL-6 (χ^2^(2) = 0.200, p = 0.905) or TNF-α (χ^2^(2) = 2.400, p = 0.301; see **Supplementary Figure 2** for individual data).

### Muscle strength

There was a significant increase in isokinetic muscle strength for leg extension at 120 degrees per second (°•s^-1^), flexion at 120°•s^-1^ and 180°•s^-1^ from PRE to POST training. Arm flexion increased at 60°•s^-1^ and 180°•s^-1^ from PRE to POST training. When normalized to lean muscle mass, only leg flexion at 120°•s^-1^ and arm flexion at 180°•s^-1^ remained significant (**Table 3)**.

**Table 3.** Absolute and normalized muscle strength values at PRE, MID and POST training

|  | PRE | MID | POST | Effect size | ANOVA/ Friedman Statistics |
| --- | --- | --- | --- | --- | --- |
| **Grip strength (Nm)** | 20.4 ± 7.1 | 20.0 ± 7.0 | 21.5 ± 7.8 | 0.12 | Greenhouse Geisser correction used, $\epsilon=0.635$, $F(1.250, 11.248)=0.312$, $p>0.05$ |
| **Normalized grip strength (Nm·kgLM<sup>-1</sup>)** | 8.3 ± 2.4 | 7.9 ± 2.0 | 8.1 ± 2.7 | -0.07 | Greenhouse Geisser correction used, $\epsilon=0.615$, $F(1.230, 11.071)=0.095$, $p>0.05$ |
| ***Elbow absolute values (Nm)*** |  |  |  |  |  |
| **Isometric extension** | 42.1 ± 10.3 | 42.8 ± 9.7 | 45.3 ± 12.0 | 0.23 | $F(2,16)=1.186$, $p>0.05$ |
| **Extension 60 °·s<sup>-1</sup>** | 31.7 ± 5.62 | 34.6 ± 7.9 | 32.3 ± 11.0 | 0.06 | Greenhouse Geisser correction used, $\epsilon=0.630$, $F(1.259, 11.334)=0.894$, $p>0.05$ |
| **Extension 120 °·s<sup>-1</sup>** | 28.8 ± 6.5 | 29.6 ± 8.5 | 28.9 ± 7.90 | 0.02 | $F(2,16)=0.126$, $p>0.05$ |
| **Extension 180 °·s<sup>-1</sup>** | 25.0 ± 3.7 | 24.7 ± 6.7 | 27.4 ± 8.7 | 0.29 | $F(2,16)=1.333$, $p>0.05$ |
| **Flexion 60 °·s<sup>-1</sup>** | 27.2 ± 8.7 | 26.5 ± 11.0<sup>+</sup> | 32.1 ± 11.2<sup>+</sup> | 0.39 | $F(2,16)=4.998$, $p<0.05$ |
| **Flexion 120 °·s<sup>-1</sup>** | 24.0 ± 9.4<sup>€</sup> | 25.2 ± 11.1<sup>€</sup> | 28.2 ± 9.2<sup>€</sup> | 0.36 | Nonparametric Friedman test, $\chi^2(2)=5.000$, $p>0.05$ |
| **Flexion 180 °·s<sup>-1</sup>** | 19.0 ± 6.8<sup>€</sup> | 22.1 ± 8.2<sup>€</sup> | 25.4 ± 9.0<sup>€</sup> | 0.66 | Nonparametric Friedman test, $\chi^2(2)=6.200$, $p<0.05$ |
| ***Elbow normalized values (Nm·kgLM<sup>-1</sup>)*** |  |  |  |  |  |
| **Isometric extension** | 17.4 ± 3.0 | 17.2 ± 3.9 | 17.1 ± 4.2 | -0.06 | $(2,18)=0.055$, $p>0.05$ |
| **Extension 60 °·s<sup>-1</sup>** | 13.2 ± 1.8 | 13.7 ± 1.8 | 12.3 ± 4.2 | -0.22 | Greenhouse Geisser correction used, $\epsilon=0.637$, $F(1.274, 11.468)=1.113$ |
| **Extension 120 °·s<sup>-1</sup>** | 11.9 ± 1.7 | 11.7 ± 2.6 | 10.9 ± 2.8 | -0.34 | $F(2,18)=1.431$, $p>0.05$ |
| **Extension 180 °·s<sup>-1</sup>** | 10.5 ± 1.9 | 9.9 ± 2.8 | 10.3 ± 2.8 | -0.08 | $F(2,18)=0.416$, $p>0.05$ |
| **Flexion 60 °·s<sup>-1</sup>** | 11.2 ± 3 | 10.3 ± 3.3 | 12.2 ± 4.3 | 0.21 | $F(2,18)=2.939$, $p>0.05$ |
| **Flexion 120 °·s<sup>-1</sup>** | 9.8 ± 2.8 | 9.9 ± 3.8 | 10.7 ± 3.6 | 0.24 | $F(2,18)=0.694$, $p>0.05$ |
| **Flexion 180 °·s<sup>-1</sup>** | 7.8 ± 2.4<sup>€</sup> | 8.8 ± 3.2*<sup>€</sup> | 9.6 ± 3.3<sup>+€</sup> | 0.49 | Nonparametric Friedman test, $\chi^2(2)=6.200$, $p<0.05$ |
| ***Knee absolute values (Nm)*** |  |  |  |  |  |
| **Extension 60 °·s<sup>-1</sup>** | 109.1 ± 29.9 | 111.7 ± 27.8 | 120.4 ± 20.6 | 0.36 | $F(2,16)=1.835$, $p>0.05$ |
| **Extension 120 °·s<sup>-1</sup>** | 87.9 ± 14.3 | 92.6 ± 19.3 | 103.5 ± 15.0* | 0.86 | Greenhouse Geisser correction used, $\epsilon=0.615$, $F(1.230, 11.071)=5.023$, $p>0.05$ |
| **Extension 180 °·s<sup>-1</sup>** | 74.2 ± 23.1<sup>+</sup> | 81.9 ± 17.7<sup>+</sup> | 85.3 ± 16.2<sup>+</sup> | 0.45 | $F(2,16)=4.817$, $p<0.05$ |
| **Flexion 60 °·s<sup>-1</sup>** | 65.3 ± 19.5<sup>€</sup> | 79.34 ± 12.2<sup>€</sup> | 74.6 ± 16.1<sup>€</sup> | 0.39 | Nonparametric Friedman test, $\chi^2(2)=4.629$, $p>0.05$ |
| **Flexion 120 °·s<sup>-1</sup>** | 57.2 ± 14.5 | 65.89 ± 16.4 | 72.22 ± 14.8* | 0.76 | $F(2,16)=6.357$, $p<0.05$ |
| **Flexion 180 °·s<sup>-1</sup>** | 45.5 ± 12.3 | 56.7 ± 15.9 | 65.9 ± 13.5*<sup>§</sup> | 1.18 | $F(2,16)=10.334$, $p<0.05$ |
| ***Knee normalized values (Nm kgLM<sup>-1</sup>)*** |  |  |  |  |  |
| **Isometric extension** | 21.4 ± 3.5 | 22 ± 4.3 | 21.8 ± 3.5 | 0.09 | $F(2, 16)=2.643$, $p>0.05$ |
| **Extension 60 °·s<sup>-1</sup>** | 15.9 ± 4.5<sup>€</sup> | 15.6 ± 3.6<sup>€</sup> | 16.5 ± 2.7<sup>€</sup> | 0.13 | Nonparametric Friedman test, $\chi^2(2)=0.6000$, $p>0.05$ |
| **Extension 120 °·s<sup>-1</sup>** | 12.8 ± 2.2<sup>€</sup> | 12.9 ± 2.5<sup>€</sup> | 14.2 ± 1.9*<sup>€</sup> | 0.53 | Nonparametric Friedman test, $\chi^2(2)=6.200$, $p<0.05$ |
| **Extension 180 °·s<sup>-1</sup>** | 10.9 ± 3.7 | 11.4 ± 2.4 | 11.7 ± 2.2 | 0.21 | $F(2,18)=0.991$, $p>0.05$ |
| **Flexion 60 °·s<sup>-1</sup>** | 9.4 ± 2.9<sup>€</sup> | 11.2 ± 2<sup>€</sup> | 9.2 ± 3.7<sup>€</sup> | -0.06 | Nonparametric Friedman test, $\chi^2(2)=3.800$, $p>0.05$ |
| **Flexion 120 °·s<sup>-1</sup>** | 8.3 ± 2.0 | 9.3 ± 2.0 | 9.0 ± 3.8 | 0.15 | Greenhouse Geisser correction used, $\epsilon=0.643$, $F(1.286, 11.578)= 0.405$, $p>0.05$ |
| **Flexion 180 °·s<sup>-1</sup>** | 7.0 ± 2.2 | 8.1 ± 2.1 | 8.2 ± 3.3 | 0.31 | $F(2.18)=0.782$, $p>0.05$ |

PRE, MID, and POST outcomes reported as mean ± standard deviation. Arm and grip strength are normalized to the lean mass (kg) of the arm performing the exercise. Leg strength is normalized to the lean mass (kg) of the leg performing the exercise. \* $P<0.05$ compared to PRE, <sup>§</sup> $P<0.05$ compared to MID, <sup>+</sup>Post-hoc test not sensitive enough to detect differences, <sup>€</sup>For non-parametric Friedman test the Bonferroni correction used for post-hoc testing resulted in significance level of $p\leq 0.017$

### Aerobic fitness

There was a significant increase in peak oxygen consumption (VO_2_ peak) and % predicted VO_2_ peak PRE to POST. We observed a trend towards an increase in VO_2_ peak normalized to lean mass. Maximum workload increased significantly from PRE to POST, even when normalized to lean mass. The submaximal data used to calculate work efficiency were only available for 8 participants, but showed a significant increase in work efficiency throughout the study (post-hoc NS, **Table 4**).

**Table 4.** Aerobic fitness variables and average daily physical activity/ sedentary time at PRE, MID and POST training

|  | PRE | MID | POST | Effect size | ANOVA/ Freidman Statistics |
| --- | --- | --- | --- | --- | --- |
| **VO<sub>2</sub> peak (L·min<sup>-1</sup>)** | 2.0 ± 0.3 | 2.2 ± 0.3 | 2.3 ± 0.2* | 0.68 | F(2, 18)=5.476, p<0.05 |
| **VO<sub>2</sub> peak (mL·kg<sup>-1</sup>·min<sup>-1</sup>)** | 36.7 ± 3.4 | 37.9 ± 4.3 | 39.5 ± 5.2 | 0.52 | F(2, 18)=1.934 |
| **VO<sub>2</sub> peak (mL·kgLM<sup>-1</sup>·min<sup>-1</sup>)** | 49.6 ± 5.7<sup>€</sup> | 50.8 ± 4.5<sup>€</sup> | 52.4 ± 4.8<sup>€</sup> | 0.42 | Nonparametric Friedman test, $\chi^2(2)=0.200$ |
| **% Predicted VO<sub>2</sub> peak** | 73.5 ± 13.9<sup>€</sup> | 76.1 ± 11.0*<sup>€</sup> | 79.3 ± 14.9*<sup>€</sup> | 0.32 | Nonparametric Friedman test, $\chi^2(2)=16.200$, p<0.05 |
| **W<sub>peak</sub> (Watts)** | 134.8 ± 19.1 | 149.0 ± 24.3* | 160.7 ± 15.6*<sup>§</sup> | 1.2 | F(2, 18)=38.589, p<0.05 |
| **W<sub>peak</sub> (Watts·kg<sup>-1</sup>)** | 2.5 ± 0.2 | 2.6 ± 0.3* | 2.8 ± 0.4* | 0.91 | F(2, 18)=12.323, p<0.05 |
| **W<sub>peak</sub> (W·kgLM<sup>-1</sup>)** | 3.3 ± 0.2 | 3.5 ± 0.3 | 3.8 ± 0.3* | 1.19 | F(2, 18)=11.606, p<0.05 |
| **% Predicted W<sub>peak</sub>** | 61.5 ± 8.1<sup>€</sup> | 66.2 ± 7.4*<sup>€</sup> | 71.6 ± 8.5*<sup>€</sup> | 0.98 | Nonparametric Friedman test, $\chi^2(2)=16.200$, p<0.05 |
| **HR<sub>peak</sub> (beats·min<sup>-1</sup>)** | 188 ± 9 | 188 ± 9 | 188 ± 10 | -0.07 | F(2, 18)=0.038, p>0.05 |
| **Work efficiency % (at PRE VO<sub>2</sub> peak)** | 18.7 ± 1.9<sup>+€</sup> | 19.4 ± 1.4<sup>+€</sup> | 19.9 ± 1.3<sup>+€</sup> | 0.49 | Nonparametric Friedman test, $\chi^2(2)=6.25$, p<0.05 |
| ***Daily physical activity and sedentary time (Min day<sup>-1</sup>)*** |  |  |  |  |  |
| **Sedentary** | 701.0 ± 52.6 | 666.8 ± 57.9 | 687.4 ± 53.8<sup>§</sup> | **Effect size** | **ANOVA Statistics** |
| **Total physical activity** | 129 ± 52.6 | 163.2 ± 57.9 | 142.1 ± 54.2<sup>§</sup> | -0.18 | Greenhouse Geisser correction used, $\epsilon=0.585$, F(1.170, 7.022)=5.827, p<0.05 |
| **Light intensity** | 91.5 ± 32.8 | 107.1 ± 33.5 | 96.3 ± 33.5<sup>§</sup> | 0.10 | F(2,12)=5.865, p<0.05 |
| **Moderate intensity** | 22.9 ± 12.1 | 28.4 ± 11.6 | 26.5 ± 11.2 | 0.22 | F(2,12)=4.487, p<0.05 |
| **Vigorous intensity** | 14.6 ± 9.1 | 27.7 ± 15* | 19.3 ± 13.1 | 0.30 | F(2,12)=1.329, p>0.05 |
| **Moderate-to-vigorous** | 37.5 ± 20.6<sup>+</sup> | 56.1 ± 26<sup>+</sup> | 45.7 ± 23.1<sup>+</sup> | 0.27 | F(2,12)=6.476, p<0.05 |
| **intensity** |  |  |  |  |  |

PRE, MID, and POST outcomes reported as mean ± standard deviation. LM= lean mass, % Predicted $W_{peak}$ based on height. \* $P<0.05$ compared to PRE, $P<0.05$ compared to MID, <sup>+</sup>Post-hoc test not sensitive enough to detect differences, <sup>6</sup>For non-parametric Freidman test the Bonferroni correction used for post-hoc testing resulted in significance level of $p\leq 0.017$

### Physical activity and sedentary time

Valid accelerometer data were available for 7 participants; 2 participants did not meet the wear time criteria at all measurement timepoints, while 1 participant did not wear the accelerometer at POST. There was a trend towards a decrease in sedentary time and an increase in total physical activity time from PRE to MID. We observed a significant increase in time spent in vigorous physical activity from PRE to MID intervention. From MID to POST, we observed a significant increase in sedentary time and a decrease in light and total physical activity (**Table 4**).

## Discussion

Overall, our 16-week aerobic and resistance exercise training program was safe, well-tolerated, and enjoyable for children and adolescents with IBD. We observed good compliance levels and adherence to supervised exercise sessions, but low adherence to home-based sessions. While there were no changes in inflammation, we did observe some improvements in body composition, muscle strength, aerobic fitness, habitual physical activity with training.

### Feasibility

In this study, 11 youth were recruited over 9 months and of these, 90.9% completed the study. A primary barrier to recruitment was the distance families had to travel to our institution for training sessions. The IBD clinic at McMaster Children’s Hospital has a large catchment area, some patients are required to travel up to 2 hours to attend the clinic. Naturally, such a far commute is not feasible 2 times a week for 16 weeks. Hence, increasing the amount of training locations or recruiting from a clinic with more local patients may increase recruitment rates.

On average, participants adhered to 77% of the prescribed exercise sessions, which did not meet our feasibility goal of at least 80% attendance. While having on-site training sessions did limit our recruitment, it is important to note that children who were enrolled had a high adherence rate to supervised sessions (87.5%), while home sessions were loosely followed with a much lower completion rate (50%). This is in accordance with previous pediatric exercise training studies that have compared adherence to supervised and unsupervised training programs.^21^ Altogether, this speaks to the need for approaches to increase motivation and the implementation of behavioural change strategies in studies designed with unsupervised training sessions.^21^ Interestingly, pediatric IBD exergame-based training studies, which also took place at home, had higher adherence rates (84.5%).^11^ Therefore, a more effective approach to increase overall program adherence may be to combine our supervised training sessions with exergame-based home sessions.

Although we were unable to report compliance for home sessions, participants were able to complete 83% of supervised sessions with no modifications to the exercise. Even in sessions requiring modification to the prescribed exercise, the modifications were minimal – often just a decrease in intensity with the participant persevering to finish the required repetitions. Overall, this suggests the training program was feasible to a certain extent, but still requires the trainer to consider the participant’s status daily. While participants wore a HR monitor for home-based sessions, the unit averaged HR over the wear period making it impossible to gauge true exercise response (i.e., resting heart rates included in the average). To improve methods of tracking home session compliance, we would suggest implementing a participant-managed training log and/or wearable device with higher resolution of data capture. Using an online platform with the log and wearable would allow real-time updates and prompts for incomplete sessions, which may be beneficial for monitoring and increasing adherence throughout the program.

### Safety

Our safety findings suggests that an increase in regular exercise does not have any significant detrimental impact on IBD-related symptoms. Such findings are in line with previous pediatric IBD and chronic inflammatory disease patients that report no significant disease exacerbation with exercise.^10,21–23^ Although two participants experienced minor flares during the study, this is not necessarily a contraindication to exercise. Given that IBD is a chronic inflammatory condition, it is expected that participants will experience sporadic exacerbation of disease symptoms regardless of what activities they engage in. Importantly, no participants withdrew from the exercise training program because of health-related concerns, and all participants remained in remission post-training. **Participant satisfaction:** Participants provided valuable feedback that can serve to inform the design of future exercise programs. For instance, the idea of individualized and goal-oriented training may help to improve motivation and adherence. There were some contradictory perspectives: one participant reported they would have been more engaged if supervised exercise sessions included their peers, while another emphasized that children with little experience being physically active may feel self-conscious and prefer the option to have one-on-one training sessions until they are comfortable training in a group. Therefore, future programs may wish to consider these options on a case-by-case basis. Furthermore, a single participant enjoyed the home-based sessions because they were more convenient and helped them recognize that they can be physically active on their own with minimal equipment. This aligns with our initial rationale for implementing home-based training sessions, and gives us hope that participants would continue to be physically active autonomously using the workouts they learned throughout the program. This is a key message that future programs may wish to emphasize to participants to increase post-training program benefits.

### Inflammatory cytokines

There were no significant changes in inflammatory cytokine levels from pre to post training. It is important to note that all of our participants were treated with drugs that modulate inflammation (e.g., corticosteroids, anti-TNF+, anti-inflammatories) throughout the study, and 7 of 10 participants changed medication dosage or type during the study. ^24^ Therefore, it is likely that any training effects on inflammation were masked by pharmacologic effects. Stratifying by medication use may be a more effective approach in future trials where the goal is to investigate the effects of regular physical activity on inflammation. Nonetheless, our results provide evidence that exercise training did not significantly increase systemic inflammation in children and adolescents treated for IBD.

### Body composition

We observed a significant increase in LBM of 2.4 ± 1.1 kg over 16 weeks of training. In healthy children of the same chronological age, we would expect to see a 1.3 ± 0.4 kg increase in LBM over a four month-period due to growth and development.^26^ This cautiously suggests that exercise training was capable of increasing lean mass in children with IBD, which is consistent with findings in children with other chronic inflammatory diseases.^25,26^ Nevertheless, without a direct control group, it is difficult to definitively conclude that this finding can be attributed to training effects or to normal growth and development.

Similarly, healthy children of the same chronological age as our participants are expected to gain ∼0.010-0.015 g•cm^-2^ of BMD per year. Our participants gained 0.012 g•cm^-2^ BMD in 16 weeks or about one third of the year.^27^ This implies that exercise may help increase BMD in children with IBD, a meaningful finding given that these youth tend to have impaired bone mass and retention.^28^ Naturally, including a non-exercising control group with IBD would provide a more definitive answer.

The reduction in FBM% we reported was likely due to an increase in overall LBM. Nonetheless, decreased FBM% and torso fat% along with an increase LBM% are encouraging, because skeletal muscle is a metabolically active tissue, which oxidizes fatty acids, even at rest.^29^ In theory, increased LBM should contribute to metabolism and energy homeostasis, and in turn, regulate fat stores and deposition as well.^28^

### Muscle strength

When normalized to extremity lean mass, only leg extension at 120°•s^-1^ and arm flexion at 180°•s^-1^ remained significant, suggesting that the increase in LBM was primarily responsible for the increase in muscle strength. Since the prescribed workouts did not expressly target isokinetic movements equivalent to 120°•s^-1^ leg extension and 180°•s^-1^ arm flexion, it is unusual that a true increase in strength would only occur for those specific angular velocities without also seeing an increase in strength tested at other angular velocities. Most of the supervised resistance training activities activated the biceps (i.e., lat pull, pec fly, seated row). However, exercises that targeted elbow extensors, like push-ups and triceps extensions, were included in the home training protocol, which was underperformed. A more balanced and targeted protocol may induce more uniform increases in strength.

### Aerobic fitness

We speculate the observed increase in absolute VO_2_ peak from pre to post was likely due to the increase in LBM, since there were no changes when VO_2_ peak was normalized to LBM. This is in contrast to previous studies in healthy children and adolescents that reported an increase in LBM-normalized VO_2_ peak following moderate intensity bi-weekly training.^30^ Significant increases in W_peak,_ even when normalized to total body mass and lean mass, were likely due to an increase in mechanical efficiency, as demonstrated by improved work efficiency at same metabolic costs. Previous studies have shown that children with IBD have lower recent hemoglobin levels, which were correlated with lower aerobic fitness.^4^ Therefore, further study may be required as to the trainability of these patients.

### Physical activity levels

We demonstrated a significant increase in vigorous physical activity from PRE to MID, and a trend towards a decrease in sedentary time and increase in total physical activity from mid to post intervention. Since data from 3 (of 10) participants were omitted from this analysis, it is possible that increasing the number of participants analyzed would strengthen the trends we observed. Furthermore, despite the strengths of device-measured physical activity, accelerometry cannot capture resistance training or stationary cycling, which were both major components of our training intervention. Exploring a combination of waist- and arm-worn activity tracking devices, or those that incorporate heart rate measurements may provide more insight into this type of structured exercise training. Importantly, our results suggest that participants were increasing their activity levels outside of the exercise intervention.

### Limitations

As previously discussed, a limitation of this pilot study is a lack of non-exercising IBD control group when evaluating the efficacy of exercise training. However, a non-exercising IBD control group was not required to achieve the primary goals of the study related to feasibility, safety, and participant feedback.

## Conclusion

The findings of this pilot study demonstrate that our 16-week exercise intervention was feasible for supervised visits and safe for youth with IBD (**Supplementary material 4**). Participants were satisfied with the training intervention, and also provided valuable feedback that can be easily implemented to improve our future training studies. From an efficacy perspective, we observed an improvement in body composition, no change in systemic inflammation, improvements in absolute aerobic fitness and work efficiency, as well as some indicators of muscle strength. Participants also demonstrated some increases in physical activity. Taken together, the findings of this pilot study are promising and should encourage further research to support the implementation of safe and effective exercise prescription as an adjunct therapy for children and adolescents with IBD.

## Data Availability

All data produced in the present study are available upon reasonable request to the authors

## Acknowledgements

The authors wish to thank the participants and their families for their time and effort. We are grateful to Ms. Chris Radoja for her tremendous support with identifying and recruiting participants. We also like to thank Mr. Peter Breithaupt for designing the exercise training intervention, as well as Ms. Jovana Milenkovic, Ms. Camilla Thorne-Tjomsland and Mr. Ian Cooper for their support with assessment and exercise training sessions. BWT was supported by a CIHR New Investigator Award.

