## Supplementary materials document for "A pilot study of exercise training for children and adolescents with inflammatory bowel disease: an evaluation of feasibility, safety, satisfaction, and efficacy"

### SUPPLEMENTARY MATERIAL

#### Supplementary material 1: Assessment visit measures

**Anthropometry:** Standing height and sitting height were measured in duplicate to the nearest 0.1 cm using a stadiometer (Harpenden Stadiometer 2109, CMS Weighing Equipment Ltd, London, UK). Sitting height and chronological age were used to estimate years from peak height velocity (PHV), which was used as a marker of biological development according to the equations of Mirwald et al (2002). Weight was measured in duplicate to the nearest 0.1 kg using a digital scale (Mott electronic scale, model LC 2424, 20-g accuracy, Santa Rosa, CA). Waist circumference (WC) was measured in triplicate 4 cm above the navel using a standard anthropometry tape. All repeated measures were averaged. Body mass index (BMI) was calculated as weight/height$^2$. Height, weight and BMI percentiles were calculated using the 2000 Centers for Disease Control growth charts and WC percentiles were calculated using reference values from 11- to 18- year-old Canadian children.[1,2] Each participant completed a self-assessment of secondary sex characteristics according to the method of Tanner – pubic hair development for boys or breast development for girls.[3] Body composition was assessed using a whole body dual energy x-ray absorptiometry (DXA) (QDR 4500A, Hologic Inc., Waltham, MA, USA) in supine position. Accompanying software (Version 12.3) was used to determine whole body and trunk fat mass (BFM, in kg and % of total body mass), lean mass (LBM, in kg and % of total body mass), bone mineral content (BMC, in kg and % of total body mass), bone mineral density (BMD, in g/cm$^2$).

**Cytokine levels:** A fasted blood sample was draw by venipuncture from the median cubital vein and collected in a 10 mL EDTA-coated tube. Plasma was isolated from blood samples using density gradient centrifugation and kept frozen until analysis. Plasma was analyzed for IL-6 and TNF-α concentrations using high sensitivity enzyme-linked immunosorbent assays (R&D Systems Inc., MN). Samples were measured in duplicate and averaged; all samples were processed in accordance with the manufacturer's protocol. Intra-assay coefficient of variation (CV) for IL-6 was 4.08%, and CV of 3.78% for TNF-α.

**Muscle strength**: An isokinetic dynamometer system (Biodex IV, Biodex, Shirley, N.Y.) was used to measure isometric and isokinetic strength of the dominant knee extensors and elbow flexors. Isometric strength testing began with a specific warm-up (3 maximal voluntary contractions). Following a short rest, participants performed three 5-s maximal voluntary contractions, each separated by a 30-s rest. For isometric knee extension testing, the knee joint was positioned at 90°. For isometric elbow flexion testing, the shoulder was positioned at 90° flexion, upper arm resting on the arm rest and the elbow at 90° flexion. Isokinetic strength was measured at three angular velocities: $60°\cdot s^{-1}$ ($1.04\ rad\cdot s^{-1}$), $120°\cdot s^{-1}$ $2.09\ rad\cdot s^{-1}$, and $180°\cdot s^{-1}$ ($3.14\ rad\cdot s^{-1}$). The range of motion of the knee was 90° (1.57 rad), starting with the knee flexed at 90° (1.57 rad) and ending in full extension. The range of motion for the elbow was 100° (1.74 rad) starting with the elbow fully extended and ending in flexion. The testing protocol began with a specific warm up (3 contractions at a progressive effort). Following a short rest, participants performed three maximal voluntary contractions at each velocity, with each velocity separated by a 3-minute rest period. The order of angular velocities was randomized. For both isometric and isokinetic strength testing, participants were instructed to contract as fast and as forcefully as possible. The highest torque was recorded in Nm and normalized to limb lean mass as measured by DXA ($Nm \cdot kg$ limb $LM^{-1}$).

**Aerobic fitness:** To assess aerobic fitness, participants completed the *McMaster All-Out Progressive Continuous Cycling Test* on an electromagnetically-braked cycle ergometer (Corival, Lode; The Netherlands). Inspired and expired gases ($O_2$ and $CO_2$) were continuously measured using a calibrated metabolic cart (Vmax29, SensorMedics, Yorba Linda, CA, U.S.A.). Heart rate (HR) was continuously measured using a Polar HR monitor (Polar Electro, Kempele, Findland). The following variables were calculated: peak oxygen uptake defined as the highest 30-sec volume of oxygen uptake ($VO_2$ peak) and expressed in L/min, ml/kg body mass/min and ml/kg LBM/min; peak mechanical power ($W_{peak}$) defined as the last workload achieved and prorated if the full 2-minute stage was not completed and expressed in watts (W), W/kg body mass and W/kg LBM; and peak HR ($HR_{peak}$) was defined as the highest heart rate achieved during the cycling test and expressed in beats/min. Percent predicted $VO_2$ and workload peak were calculated using equations created from a repository of aerobic fitness tests results preformed in our laboratory. Work efficiency was also calculated based on the equation described by Garby.[4]

**Max repetitions in 45 seconds:** For bodyweight exercises, participants were asked to repeat the exercise as many times as they could in 45 seconds. Participants were times with a chronometer and the number of well-performed repetitions was recorded. Max repetition in 45 seconds testing was performed at the start of the training program and repeated at mid-training to adjust workloads.

**1RM:** For each resistance exercise, participants were first asked to lift a given load. Following the lift, participants were asked how their body felt while performing the exercise on a scale from 0 to 10 (0=extremely easy, 10=cannot lift more than once).[5] The load was increased by 1-5 kg until a perceived difficulty of 10 was reached, which indicated a participant's 1-RM for a given exercise. The increments were dependent on the effort required for the lift and got progressively smaller as the participant approached the 1-RM.[6] 1-RM was achieved within 6-10 repetitions, and used to standardize the training sessions. 1-RM testing was performed at the start of the training program and repeated at mid-training to adjust workloads.

**Habitual physical activity:** At the end of each assessment session (pre, mid, post), participants were given an ActiGraph GT3X accelerometer (ActiGraph LLC, Pensacola, FL) and instructed to wear the device around their waist over their right hip, during all waking hours for 7 days, with the exception of water activities. Participants were asked to complete a logbook to indicate any times the device was removed. When the device was returned, all data were downloaded in 3-sec intervals, cleaned and processed using ActiLife software. Only participants with at least 3 valid days of accelerometer wear per assessment period, defined as wearing the device for at least 10 hours per day, were included in the final analysis. Evenson cut points to quantify time spent in being sedentary, as well as light, moderate, and vigorous intensity physical activity.[7] Data are reported as average minutes per day, normalized to average daily wear time of all participants for all sessions.

**Supplementary material 2: Exercise intervention details**

Each week participants completed 2 supervised training sessions with a trainer at McMaster University and 1 training session independently at home. Each supervised and home training session consisted of resistance training and aerobic exercises. A sample training week is provided in Supplementary table 1.

**Supervised training sessions**

*Resistance training*

At McMaster, the intensity of machine-based resistance exercises began at 40% of participants' baseline 1-RM, while body weight exercises began at 40% of maximum repetitions completed in 45-sec. As the training program progressed, the resistance exercise stimulus was increased in a step-wise fashion by alternating between increasing the intensity of exercise and the number of repetitions prescribed. After 8 weeks of training, 1-RM and 45-sec max were re-assessed and new values were used to calculate subsequent workout intensities.

*Aerobic exercises*

The intensity of aerobic exercise was based on the participant's peak workload ($W_{peak}$ ) from the aerobic fitness test. To begin, participants performed continuous cycling for 3 minutes at 50% $W_{peak}$, followed by 30 sec high intensity intervals at 80% $W_{peak}$ separated by a 30 sec rest interval at 50% $W_{peak}$. Finally participants completed steady state cycling for 10 min at 40% of the participant's baseline $W_{peak}$. As the training program progressed, the aerobic exercise stimulus was increased in a step-wise fashion by alternating between increasing duration and workload. After 8 weeks of training, $W_{peak}$ was re-assessed and new values were used to calculate subsequent cycling intensities. HR was monitored continuously throughout the training sessions using a Polar HR monitor (Polar Electro, Kempele, Findland).

**Home-based sessions**

To ensure proper understanding of the home-based exercise protocol, one researcher (RGW) taught the participants each exercise, and provided them with a manual containing pictures and descriptions of the various exercises. Similar to the supervised sessions, considerations were made to ensure a continuous progression in the exercise stimulus during the home sessions. Participants were provided with a HR monitor to wear during home sessions to allow them to monitor exercise intensity.

Aerobic exercise included 30 sec intervals beginning at 80% of baseline $HR_{peak}$ and steady state aerobic exercise beganning with 5 min at 40% of baseline $HR_{peak}$. Bodyweight exercises began at 40% of maximum repetitions complete in 45 sec. After 8 weeks of training, $HR_{peak}$ and max reps in 45 sec were re-assessed and new values were used to calculate subsequent intensities. During each training session, the HR monitor recorded data, which was then uploaded to an online training log by an investigator. To encourage the completion of home sessions, the investigator contacted participants and/or their families weekly by e-mail, phone call or text message.

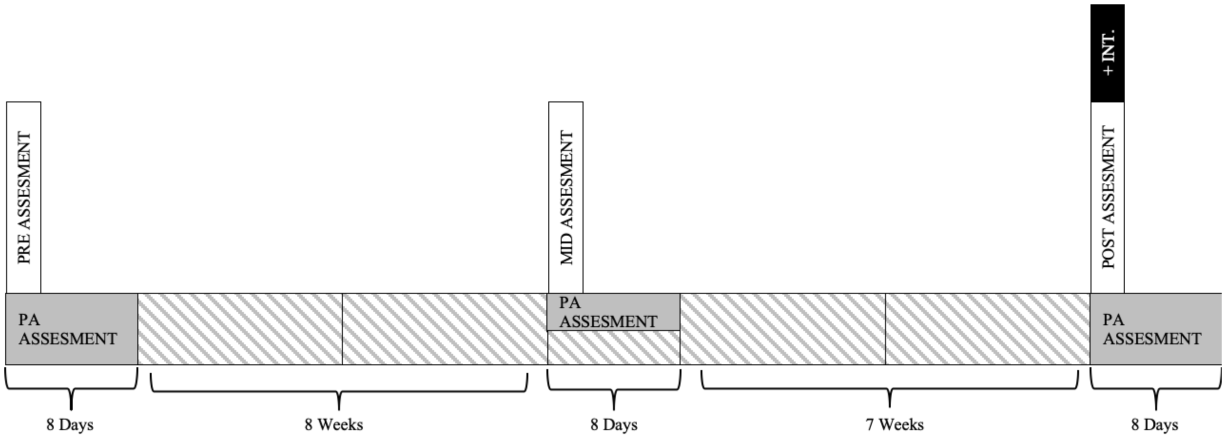


**Supplementary Figure 1.** Schematic of the study outline. After each assessment session, participants were given an accelerometer to wear for 7 consecutive days. For more information see "assessment visits" above. Striped bars indicate training time. PA Assessment= participant is wearing an accelerometer for PA monitoring, INT.= participant satisfaction interview.

**Supplementary Table 1.** Training protocol

| **McMaster University session protocol** | | | | **Home session protocol** | |
|---|---|---|---|---|---|
| **DAY 1** | | **DAY 2** | | **1 X/ WEEK** | |
| **5 min** | **WARM UP** – cycle | **5 min** | **WARM UP** – cycle | **5 min** | **WARM UP** – *skipping or activity of choice* |
| **1-5 min** | **AEROBIC**<br>Cycle – *Intervals*<br>Start: 3 min @ 50% $W_{peak}$<br>Intervals: 1-5 x 30s @ 80-100% $W_{peak}$, each followed by 30s @ 50% $W_{peak}$<br>End: 3 min @ 50% $W_{peak}$ | **1-5 min** | **AEROBIC**<br>Cycle – *Intervals*<br>Start: 3 min @ 50% $W_{peak}$<br>Intervals: 1-5 x 30s @ 80-100% $W_{peak}$, each followed by 30s @ 50% $W_{peak}$<br>End: 3 min @ 50% $W_{peak}$ | **1-5 min** | **AEROBIC**<br>Skipping or activity of choice – *Intervals*<br>*1-5 x 30s @ 80-100%* $HR_{peak}$, each followed by 30s rest |
| **10-20 min** | **RT**<br>Leg Press<br>Chest Press<br>Lat Pull Down<br>Low Back Extension<br>Bicycle<br>*1-3 sets, 12-15 reps, 40-70% 1-RM or max reps in 45s* | **10-20 min** | **RT**<br>Leg Extension<br>Leg Curl<br>Pec Fly<br>Seated Row<br>Abdominal Crunches<br>*1-3 sets, 12-15 reps, 40-70% 1-RM or max reps in 45s* | **10-20 min** | **RT**<br>Push Ups<br>Wall Squats<br>Band Rows<br>Band Bicep Curls<br>Band Tricep Extensions<br>Plank<br>*1-3 sets, 40-70% of max reps in 45s* |
| **10-20 min** | **AEROBIC**<br>Cycle – *Steady State* | **10-20 min** | **AEROBIC**<br>Cycle – *Steady State* | **5-15 min** | **AEROBIC**<br>Activity of choice – *Steady State* |

| | | | | | |
|---|---|---|---|---|---|
| | 40-70% $W_{peak}$ | | 40-70% $W_{peak}$ | | 40-70% $HR_{peak}$ |
| **5 min** | **COOL DOWN** *– dynamic and static stretching* | **5 min** | **COOL DOWN** *– dynamic and static stretching* | **5 min** | **COOL DOWN –** *dynamic and static stretching* |

RT, resistance training. HR, heart rate. W, workload. Ranges in time, intensities and number of sets and repetitions reflect increases in the training stimulus over the course of the 16-week exercise training program.

**Supplementary table 2.** Participant feedback on training program likes and dislikes

| **Likes** | **Frequency reported** |
|---|---|
| Resistance training machines | 6 |
| Skipping exercises | 1 |
| Biking | 1 |
| Stretching | 1 |
| Exercise variety | 1 |
| Home sessions | 1 |
| McMaster training environment | 2 |
| BOOST drink | 1 |
| Positive feeling | 1 |
| Getting stronger | 4 |
| **Dislikes** | |
| Biking | 6 |
| Leg extensions | 2 |
| BOOST drink | 1 |
| Home sessions | 1 |
| Seated row | 1 |

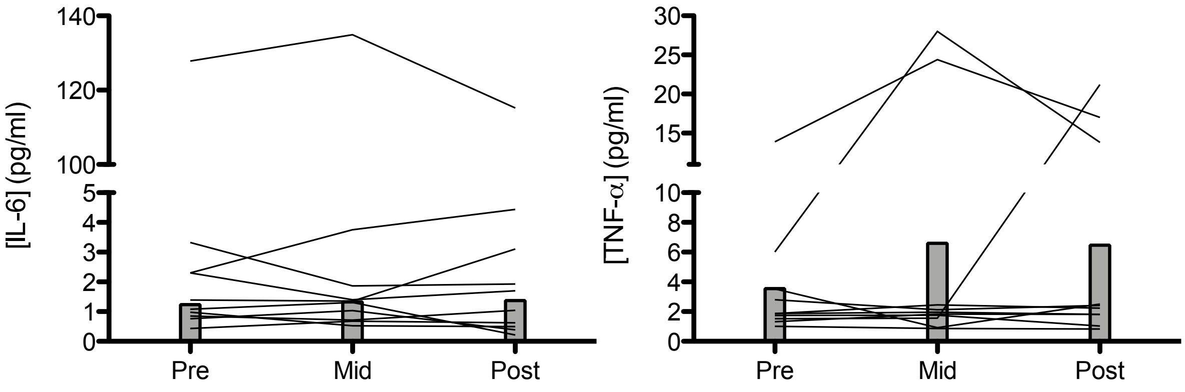


**Supplementary Figure 2.** Circulating cytokine levels. The grey lines denote data for individual participants at each time point. The grey bars demonstrate the group median at each time point. A single participant, IBDM-09, had abnormally high levels of IL-6 (>110 pg/mL) for all three assessment points; however, Freidman test results were unchanged when this participant was excluded.

Supplementary Table 4. Average number of stools a day during the study

| NUMBER OF STOOLS A DAY | M1W1 | M2W2 | M2W3 | M3W3 | M4W1 | M4W3 |
|---|---|---|---|---|---|---|
| IBDM01 | 3 | 3 | 2 | 3 | 4 | 4 |
| IBDM02 | 1 | 1 | 1 | 1 | 1 | 1 |
| IBDM03 | 1.5 | 1.5 | 1.5 | 1.5 | 1.5 | N/A |
| IBDF04 | 2 | 4 | 2 | 2 | 2 | 2 |
| IBDM05 | N/A* | N/A* | N/A* | N/A* | N/A* | N/A* |
| IBDM06 | N/A* | N/A* | N/A* | N/A* | N/A* | N/A* |
| IBDM07 | 1 | 2 | 2 | | 2 | 2 |
| IBDM08 | 2 | 3 | 2 | 3 | 3 | 3 |
| IBDM09 | N/A* | N/A* | N/A* | N/A* | N/A* | N/A* |
| IBDM10 | 1 | 1 | 1 | 1 | N/A | N/A |

*Patient has an ostomy pouch and cannot answer this question

Supplementary Table 5. Stool consistency during the study

| STOOL CONSISTENCY | M1W1 | M2W2 | M2W3 | M3W3 | M4W1 | M4W3 |
|---|---|---|---|---|---|---|
| IBDM01 | Formed | Formed | Formed | Formed | Formed | Semi formed |
| IBDM02 | Formed | Formed | Formed | Formed | Formed | Formed |
| IBDM03 | Formed | Formed | Formed | Formed | Formed | N/A |
| IBDF04 | Semi formed | Liquid | Formed | Formed | Formed | Formed |
| IBDM05 | N/A* | N/A* | N/A* | N/A* | N/A* | N/A* |
| IBDM06 | N/A* | N/A* | N/A* | N/A* | N/A* | N/A* |
| IBDM07 | Formed | Formed | Formed | Formed | Formed | Formed |
| IBDM08 | Semi formed | Semi formed | Semi formed | Formed | Semi formed | Semi formed |
| IBDM09 | N/A* | N/A* | N/A* | N/A* | N/A* | N/A* |
| IBDM10 | Formed | Formed | Formed | Formed | N/A | N/A |

*Patient has an ostomy pouch and cannot answer this question

Supplementary Table 6. Blood in stool during the study

| BLOOD IN STOOL | M1W1 | M2W2 | M2W3 | M3W3 | M4W1 | M4W3 |
|---|---|---|---|---|---|---|
| IBDM01 | None | None | None | None | None | None |
| IBDM02 | None | Trace | None | None | None | None |
| IBDM03 | None | None | Trace | None | None | N/A |
| IBDF04 | None | None | None | None | None | None |
| IBDM05 | N/A* | N/A* | N/A* | N/A* | N/A* | N/A* |
| IBDM06 | N/A* | N/A* | N/A* | N/A* | N/A* | N/A* |
| IBDM07 | None | None | None | None | None | None |
| IBDM08 | None | None | None | None | None | None |
| IBDM09 | N/A* | N/A* | N/A* | N/A* | N/A* | N/A* |
| IBDM10 | None | None | None | None | N/A | N/A |

*Patient has an ostomy pouch and cannot answer this question

Supplementary Table 7. Mucus in stool during the study

| MUCUS IN STOOLS | M1W1 | M2W2 | M2W3 | M3W3 | M4W1 | M4W3 |
|---|---|---|---|---|---|---|
| IBDM01 | None | None | None | None | None | Trace |
| IBDM02 | None | None | None | None | None | None |
| IBDM03 | None | None | None | None | None | N/A |
| IBDF04 | None | Trace | None | None | None | None |
| IBDM05 | N/A* | N/A* | N/A* | N/A* | N/A* | N/A* |
| IBDM06 | N/A* | N/A* | N/A* | N/A* | N/A* | N/A* |
| IBDM07 | None | None | None | None | None | None |
| IBDM08 | None | None | None | None | None | None |
| IBDM09 | N/A* | N/A* | N/A* | N/A* | N/A* | N/A* |
| IBDM10 | None | None | None | None | N/A | N/A |

*Patient has an ostomy pouch and cannot answer this question

Supplementary Table 8. Perianal discomfort during the study

| **PERIANAL DISCOMFORT** | **M1W1** | **M2W2** | **M2W3** | **M3W3** | **M4W1** | **M4W3** |
|---|---|---|---|---|---|---|
| IBDM01 | No | No | N/A | No | No | No |
| IBDM02 | No | No | No | No | No | No |
| IBDM03 | N/A | N/A | No | No | No | N/A |
| IBDF04 | No | No | N/A | No | No | No |
| IBDM05 | N/A* | N/A* | N/A* | N/A* | N/A* | N/A* |
| IBDM06 | N/A* | N/A* | N/A* | N/A* | N/A* | N/A* |
| IBDM07 | No | No | No | No | No | No |
| IBDM08 | No | No | Yes | Yes | No | No |
| IBDM09 | N/A* | N/A* | N/A* | N/A* | N/A* | N/A* |
| IBDM10 | No | No | No | No | N/A | N/A |

Supplementary Table 9. Nausea during the study

| **NAUSEA** | **M1W1** | **M2W2** | **M2W3** | **M3W3** | **M4W1** | **M4W3** |
|---|---|---|---|---|---|---|
| IBDM01 | No | No | No | No | No | No |
| IBDM02 | No | Yes | No | No | No | No |
| IBDM03 | No | No | No | No | No | N/A |
| IBDF04 | No | Yes | No | No | No | No |
| IBDM05 | Yes | No | No | No | No | No |
| IBDM06 | No | No | No | No | No | No |
| IBDM07 | No | No | No | No | No | No |
| IBDM08 | No | No | No | No | No | No |
| IBDM09 | No | No | No | No | No | No |
| IBDM10 | Yes | Yes | Yes | No | N/A | N/A |

Supplementary Table 10. Vomiting during the study

| VOMITING | M1W1 | M2W2 | M2W3 | M3W3 | M4W1 | M4W3 |
|---|---|---|---|---|---|---|
| IBDM01 | No | No | No | No | No | No |
| IBDM02 | No | Yes | No | No | No | No |
| IBDM03 | No | No | No | No | No | N/A |
| IBDF04 | No | Yes | No | No | No | No |
| IBDM05 | No | No | No | No | No | No |
| IBDM06 | No | No | No | No | No | No |
| IBDM07 | No | No | No | No | No | No |
| IBDM08 | No | No | No | No | No | No |
| IBDM09 | No | No | No | No | No | No |
| IBDM10 | No | No | No | No | N/A | N/A |

Supplementary Table 11. Extraintestinal manifestations during the study

| EXTRAINTESTINAL MANIFESTATIONS | M1W1 | M2W2 | M2W3 | M3W3 | M4W1 | M4W3 |
|---|---|---|---|---|---|---|
| IBDM01 | None | None | None | None | None | None |
| IBDM02 | None | None | None | None | None | None |
| IBDM03 | None | N/A | None | None | None | N/A |
| IBDF04 | None | N/A | None | None | None | None |
| IBDM05 | None | None | None | None | Knee pain | None |
| IBDM06 | None | None | None | None | None | None |
| IBDM07 | None | None | None | None | None | None |
| IBDM08 | None | None | None | None | None | None |
| IBDM09 | None | None | None | None | None | None |
| IBDM10 | Oral ulcers | Oral ulcers | Oral ulcers | None | N/A | N/A |

Supplementary Table 12. Nutrition during the study

| **NUTRITION** | **M1W1** | **M2W2** | **M2W3** | **M3W3** | **M4W1** | **M4W3** |
|---|---|---|---|---|---|---|
| IBDM01 | Good | Good | Good | Good | Good | Good |
| IBDM02 | Good | Good | Good | Good | Good | Good |
| IBDM03 | Good | Good | Good | Good | Good | N/A |
| IBDF04 | Good | | | Good | Some wasting | Some wasting |
| IBDM05 | Good | Good | Good | Good | Good | Good |
| IBDM06 | Some wasting | Some wasting | Good | Some wasting | Some wasting | Some wasting |
| IBDM07 | N/A | Some wasting | Good | Some wasting | Some wasting | Some wasting |
| IBDM08 | Good | Good | Good | Good | Good | Good |
| IBDM09 | Good | Good | Good | Good | Good | Good |
| IBDM10 | Some wasting | Some wasting | Some wasting | Good | N/A | N/A |

Supplementary Table 13. Appetite during the study

| **APPETITE** | **M1W1** | **M2W2** | **M2W3** | **M3W3** | **M4W1** | **M4W3** |
|---|---|---|---|---|---|---|
| IBDM01 | Normal | Normal | Normal | Normal | Normal | Normal |
| IBDM02 | Normal | Normal | Normal | Normal | Normal | Normal |
| IBDM03 | Normal | Normal | Normal | Normal | Normal | N/A |
| IBDF04 | Normal | N/A | N/A | Normal | Improved | N/A |
| IBDM05 | Improved | Improved | Improved | Improved | Normal | Improved |
| IBDM06 | Normal | Normal | Normal | Normal | Normal | Improved |
| IBDM07 | N/A | Normal | Normal | Normal | Normal | Decreased |
| IBDM08 | Normal | Normal | Normal | Normal | Normal | Normal |
| IBDM09 | Normal | Normal | Normal | Normal | Normal | Normal |
| IBDM10 | Decreased | Improved | Improved | Improved | N/A | N/A |

Supplementary Table 14. Supplements taken during the study

| SUPPLEMENTS | M1W1 | M2W2 | M2W3 | M3W3 | M4W1 | M4W3 |
|---|---|---|---|---|---|---|
| IBDM01 | Vitamin D<br>Iron<br>Calcium | Vitamin D<br>Iron<br>Calcium | Vitamin D,<br>Iron<br>Calcium | Iron<br>Calcium | Vitamin D | Vitamin D |
| IBDM02 | No | No | No | No | B-12 | No |
| IBDM03 | No | N/A | No | N/A | No | N/A |
| IBDF04 | No | N/A | N/A | Omega-3 | N/A | N/A |
| IBDM05 | No | N/A | N/A | No | No | No |
| IBDM06 | No | No | No | No | No | No |
| IBDM07 | N/A | No | No | No | No | No |
| IBDM08 | Probiotics<br>Multivitamins | Probiotics<br>Multivitamins | Probiotics<br>Multivitamins | Probiotics<br>Multivitamins | Probiotics<br>Multivitamins | Probiotics<br>Multivitamins |
| IBDM09 | No | Iron, Zoloft | No | No | No | No |
| IBDM10 | No | No | No | Vitamin C, D | N/A | N/A |

Supplementary Table 15. Use of external feeds during the study

| EXTERNAL FEEDS | M1W1 | M2W2 | M2W3 | M3W3 | M4W1 | M4W3 |
|---|---|---|---|---|---|---|
| IBDM01 | N/A | N/A | N/A | No | No | No |
| IBDM02 | No | No | No | No | No | No |
| IBDM03 | N/A | N/A | No | N/A | No | N/A |
| IBDF04 | N/A | N/A | N/A | No | N/A | N/A |
| IBDM05 | N/A | N/A | N/A | No | No | No |
| IBDM06 | No | N/A | No | No | No | No |
| IBDM07 | N/A | No | No | No | No | No |
| IBDM08 | N/A | N/A | No | No | No | No |
| IBDM09 | No | N/A | No | No | No | No |
| IBDM10 | No | No | No | No | N/A | N/A |

Supplementary Table 16. Menstrual status during the study

| MENSTRUAL CYCLE | M1W1 | M2W2 | M2W3 | M3W3 | M4W1 | M4W3 |
|---|---|---|---|---|---|---|
| IBDF04 | Regular | Regular | Regular | Regular | Regular | Regular |

All other participants in this study were male.
